# Association of Deep Learning-Derived Histologic Features of Placental Chorionic Villi with Maternal and Infant Characteristics in the New Hampshire Birth Cohort Study

**DOI:** 10.1101/2025.04.22.25325465

**Authors:** Elizabeth C. Anderson, Gokul Srinivasan, Caitlin G. Howe, Edward Zhang, Catherine Jeon, Gnan Suchir Gupta Paruchuri, Leah Zhang, Lindsay Hwang, Aditya Sengar, Neha Reddy, Anmol Karan, Andrew Chen, Julia Shen, Onyinyechi Owo, ZoëFaith Caraballo-Bobea, Camilo Khatchikian, Thomas J. Palys, Louis J. Vaickus, Juliete C. Madan, Margaret R. Karagas, Jessica L. Bentz, Joshua J. Levy

## Abstract

**Introduction:** Quantification of placental histopathological structures is challenging due to a limited number of perinatal pathologists, constrained resources, and subjective assessments prone to variability. Objective standardization of placental structure is crucial for easing the burden on pathologists, gaining deeper insights into placental growth and adaptation, and ultimately improving maternal and fetal health outcomes.

**Methods:** Leveraging advancements in deep-learning segmentation, we developed an automated approach to detect over 9 million placenta chorionic villi from 1,531 term placental whole slide images from the New Hampshire Birth Cohort Study. Using unsupervised clustering, we successfully identified biologically relevant villi subtypes that align with previously reported classifications – terminal, mature intermediate, and immature intermediate – demonstrating consistent size distributions and comparable abundance. We additionally defined tertile-based combinations of villi area and circularity to characterize villous geometry. This study applies these cutting-edge AI methods to quantify villi features and examine their association with maternal and infant characteristics, including gestational age at delivery, maternal age, and infant sex.

**Results:** Increasing gestational age at delivery was statistically significantly associated (p=0.003) with an increase in the proportion of mature intermediate villi and a decrease in the proportion of the smallest, most circular villi (p < 0.001). Maternal age and infant sex were not statistically significantly associated with measures of villous geometry.

**Discussion:** This work presents a workflow that objectively standardizes chorionic villi subtypes and geometry to enhance understanding of placental structure and function, while providing insights into the efficiency, growth, and the architecture of term placentas which can be used to inform future clinical care.

## Introduction

Abnormal placental development, including delayed or accelerated maturation, can lead to complications including fetal intrauterine growth restriction, preterm birth, stillbirth, preeclampsia, as well as future maternal cardiovascular disease and metabolic disorders in the offspring.^1–3^ Accurate diagnosis of abnormal placental development can provide retrospective insight into the *in utero* environment, guide future pregnancy care, and inform long-term child health.^4,5^ Establishing objective measures of placental maturation is essential for optimizing clinical care and improving perinatal outcomes.

Chorionic villi are the primary structural and functional units of the placenta and deviations in their size or morphology may reflect abnormalities in their development, which may have significant implications for fetal growth and development. Villi are finger-like projections of placental tissue in the form of tree-like vasculature, and they create a physical barrier between maternal and fetal circulation and regulate selective transport between the mother and fetus. Villi subtypes include mesenchymal, stem, mature intermediate, immature intermediate, and terminal (**Figure S1, Table S1**).^6–8^ By term, terminal villi are the main site of bi-directional exchange between the mother and fetus.^9^ Reportedly, terminal villi constitute the largest proportion of villi in term placentas, followed by mature intermediate, stem, and immature intermediate villi.^6,10–16^ Additionally, villi shape across all subtypes is reported to remain relatively stable through gestation^17^ though there are reported variations based on maternal age but not fetal sex.^8,18,19^

A limited number of studies have aimed to quantify villi structure and geometry; however, conflicting methods make comparisons between studies complicated.^14,20,21^ More reliable and quantitative descriptions of placental growth are hindered by the shortage of pathologists specializing in placental histopathology, biobanking logistics such as sample preservation and storage, variation in placental terminology, subtle differences in histopathological features, and the sheer quantity of villi in each placenta sample.^22^ Additionally, even among trained experts in perinatal pathology, concordance of histopathological examination, including segmentation of chorionic villi, is poor.^23,24^ Thus, there has been a recent call for increased standardization across placental pathologists, which includes a standard process for classifying placentas into normal, advanced, and delayed maturation.^5^

Automated approaches, such as artificial intelligence (AI) technologies, hold promise as valuable tools to enhance histopathological examination in a standard and reproducible manner. AI application has already demonstrated success in several end-to-end approaches.^25^ For example, Mobadersany et al., reported a tool to predict the gestational age of 154 WSIs with an r^2^ of 0.8859.^26^ Kidron et al. leveraged the open-source ImageJ analysis tool for the automated segmentation and analysis of placental villi across various maturation subtypes, reporting substantial differences in villi count and size.^27^ Salsabili et al. published the results of an image segmentation pipeline for the detection of placental villi, achieving accuracies of approximately 82% when validated across placentas from normal and preeclamptic patients.^28^

Despite progress, AI-based methods remain impractical for clinical use due to their complexity, variability, and dependence on subjective assessments prone to inter-observer differences and various interpretations. Many approaches rely on image heuristics like boundary detection and color thresholding, which limit generalizability across diverse placental samples and imaging protocols, and are constrained by time, cost, and small image regions.^28,29^ While some deep learning methods now summarize features across slides, they don’t quantify specific villi features. Standardized villi quantification could improve understanding of placental development and pregnancy-related diseases the identification of abnormalities and improving insights into pregnancy-related diseases that impact maternal and fetal health, an area that remains understudied.

To address these limitations, we aimed to (1) develop a rapid and scalable annotation and segmentation algorithm that leverages advancements in AI to improve the detection and characterization of chorionic villi, (2) quantify villous subtypes and their size and circularity, and (3) evaluate if quantitative metrics of villi are associated with maternal or infant characteristics that have been highlighted in prior literature, including gestational age at delivery, maternal age, and infant sex. To accomplish these aims, we have digitized over 1,500 placentas from the New Hampshire Birth Cohort Study (NHBCS).^30^

## Methods

### Study Sample, Placenta Collection and Digitization

The NHBCS is an ongoing, longitudinal pregnancy cohort based in rural New Hampshire that has enrolled over 3,000 mother-infant dyads. Current eligibility criteria include English literacy, a singleton pregnancy, and no intention or plans to relocate. This analysis used demographic information from participants between January 2009 and December 2022.

Gestational age at delivery, maternal age, and infant sex were selected from the NHBCS as primary exposures, guided by recommendations from J.B. (a board-certified pathologist) and informed by prior literature. Specifically, while gestational age at delivery and maternal age have previously been associated with variations in villi geometry and growth, infant sex has not and served as a negative control.^8,18–21,31^ Each characteristic (gestational age at delivery, maternal age, and infant sex) is clinically relevant to placental development and can potentially influence fetal growth and pregnancy outcomes. Gestational age at delivery was derived from electronic medical records, using ultrasound measurements taken near 13 weeks gestation when available, or estimated from the expected date of confinement. When neither was available or logical, gestational age at delivery was estimated based on the last menstrual period. Gestational age at delivery was evaluated continuously and categorically (i.e., preterm (≤37 weeks) vs. term (≥37 weeks)). Maternal age at delivery was derived from delivery medical records and infant sex was abstracted from electronic medical records.

Placentas were collected from NHBCS participants upon delivery. A section of the fetal portion of each placenta, approximately 1 cm deep and 1-2 cm across, was excised near the base of the umbilical cord insertion. These biopsies were transported to the Dartmouth Hitchcock Department of Pathology via the hospital’s pneumatic tube system and stored at 4°C and fixed in 10% neutral-buffered formalin for at least 24 hours. The formalin-fixed biopsies were then paraffin embedded and Hematoxylin and eosin (H&E) stained, imaged at 40X resolution using Leica Aperio GT450 image scanners housed in the Pathology Shared Resource.

Whole slide images (WSIs) from n=1,531 NHBCS participants’ placentas were digitized, and n=1,348 were included in the analytic subset after exclusions were applied, as detailed in **Figure S2**. In this analytic subset, the median BMI was 24.8 kg/m², and 58.5% of participants held a college degree or higher (**Table 1**). The median (IQR) gestational age at delivery was 39.3 weeks (range: 30.9 - 43.1 weeks). Most participants (92.3%) delivered at ≥37 weeks of gestation. The median (IQR) maternal age was 32.1 (28.8-35.2) years. The majority of participants (73.6%) were under 35 years, the cutoff for advanced maternal age, which is associated with declining fertility and higher risks of genetic abnormalities and pregnancy complications.^32^ The overlap in participants missing gestational age, maternal age, and infant sex was high (62.9%) (**Figure S3**).

**Table 1:** Population Characteristics.

| Characteristic |  | n=1,348 NHBCS Participants |
| --- | --- | --- |
| Maternal age, years |  | 32.08 (28.82 - 35.20) |
| Maternal age <35 years |  | 930 (73.6%) |
| Gestational age at delivery, weeks |  | 39.29 (38.43 - 40.29) |
| Preterm, delivery at <37 weeks gestation |  | 96 (7.63%) 3 |
| Maternal pre-pregnancy BMI, kg/m<sup>2</sup> |  | 24.80 (22.24 - 29.29) |
| Placenta weight, grams |  | 580.00 (490.00 - 660.00) |
| Female Sex |  | 598 (48.9%) |
| Education | Up to any college | 253 (18.7%) |
|  | College graduate or higher | 790 (58.5%) |
|  | Missing | 307 (22.7%) |
| Delivery Type | Vaginal | 887 (65.7%) |
|  | C-section | 362 (26.8%) |
|  | Missing | 101 (7.5%) |
| Parity | 0 | 570 (42.2%) |
|  | 1 | 486 (36%) |
|  | 2 | 172 (12.7%) |
|  | ≥3 | 57 (4.2%) |
|  | Missing | 65 (4.8%) |
| Smoking | Smoked while pregnant | 59 (4.4%) |
|  | Ever Smoked (but not during pregnancy) | 47 (3.5%) |
|  | Second-hand smoking exposure during pregnancy | 43 (3.2%) |
|  | No reported smoking or second-hand smoking exposure | 1,086 (80.4%) |
|  | Missing | 115 (8.5%) |

### Whole Slide Image Annotation of Villous Structures

Whole slide images (WSIs) were annotated using QuPath,^33^ with most villi contours generated using the Segment Anything Model (SAM)^34^ plugin to accelerate the process while maintaining precision. Each annotation was reviewed at least twice by E.C.A and standardized by J.B. to ensure consistency. A total of 4,975 villi were annotated from 50 regions of interest. These regions were used to generate training and validation data via a non-overlapping sliding window approach with 1024×1024-pixel patches. A detailed description of these methods is included in the **Supplemental Text** under “WSI Annotation.”

### Post-Processing, Modeling, and Validation

Image segmentation models considered for this project were Mask R-CNN^35^ and YOLOv8^36^ for the complementary tasks of object detection and instance segmentation. These were selected due to their active maintenance, widespread usage, and generally low training and inference costs. Their specific training protocols, configurations, post-processing, performance evaluation, and validation are described in the supplementary materials (**Supplemental Text,** “Instance Segmentation Model Details”).

To briefly summarize post-processing methods, a sliding window with overlapping patches was used to address villi truncation at patch edges, and duplicate detections were merged using a connected components graph based on an IoU threshold of 0.05. Model performance was evaluated via five-fold cross-validation using segmentation mean average precision (mAP50 and mAP50-95) and validated by correlating predicted and ground truth villi counts using Pearson correlation. The final model was applied to the remaining WSIs, with results saved in geojson format and reviewed by J.B. for quality assurance.

### Villi Subtype Clustering

To separate the annotations into interpretable, biologically relevant groups, K-means clustering was conducted on a random subset of 150 annotated villi per WSI (n=222,319 total) using features from a pre-trained VGG16^37^ model and villous dimensions, followed by dimensionality reduction with UMAP. Clusters were identified based on visual inspection, and artifact clusters containing non-villous structures were excluded along with additional annotation artifacts filtered using a custom perimeter-based algorithm. Final cluster proportions were calculated for each WSI based on the number of remaining valid villi per cluster. A detailed explanation of the villi clustering methods is included in the **Supplemental Text**, “Villi Clustering.”

### Villi Tertile Proportions

To better understand the geometry of placental villi, we sought to assess their raw circularity and size independently of villi subtype cluster assignments. Although the clustering algorithm may reveal distinct subtypes of villi (i.e., immature, mature, and terminal villi), the absence of clear visual boundaries between clusters prompted evaluation of basic villi characteristics. By categorizing each villi into a size tertile (“S,” between 1 and 3) and a circularity ratio tertile (“C,” between 1 and 3), based on global measures across all WSIs, we aimed to identify simpler metrics that might serve as biomarkers of villous maturation. Further details about the specific calculation are included in **Supplemental Text,** “Tertile Classification Creation.” This approach resulted in nine possible combinations of tertile size and circularities (S1:C1 - S3:C3), and for each WSI we calculated the count and proportion of villi belonging to each combination to capture the distribution of villous characteristics, referred to as *tertile proportions*.

### Maternal and Infant Characteristics Associations

Gestational age at delivery was examined continuously (independent variable) using two statistical models. The first model assessed villi subtype proportion (dependent variable) from the clustering algorithm using a beta regression model (*betareg* package, R v4.2.2).^38^ The second model examined villi size and circularity using tertile proportions in each WSI (dependent variable) also using a beta regression. Additionally, we aimed to directly compare the changes in the largest, most irregular shaped villi (S3:C1) with the smallest, roundest villi (S1:C3) across gestational age at delivery. To achieve this, we used a binomial logistic regression model, using the “glm” function in R. This approach modeled the total number of S3:C1 villi (𝑦_!_) within a slide through *y_i_*∼*Binom*(*n_i_*, *p_i_*); *logit*(*p_i_*) = 𝛽_0_ + 𝛽_1_ *GA_i_*, where *𝑛_i_* is the number of S3:C1 and S1:C3 villi and *p_i_* is the estimated proportion of S3:C1 villi. This model assessed the log-odds of the change in the proportion of these two villi types as a function of gestational age at delivery.

In supplemental analyses, the effect of preterm status on villi subtype proportions was evaluated with a binomial piecewise regression (“glm” function from “stats” package in R) given known structural and transcriptomic differences between term and preterm placentas.^39,40^ The primary outcomes were the proportions of intermediate (mature and immature) to terminal villi, with gestational age and preterm status as independent variables. Interaction terms were included to assess whether the relationship between gestational age and villi subtype varied by preterm status. Odds ratios were calculated from exponentiated model coefficients, and statistical significance was evaluated using a p-value threshold of 0.05 and 95% confidence intervals. An odds ratio greater than one indicates a positive association between gestational age at delivery and villi subtype/tertile proportion.

In addition to gestational age at delivery, maternal age and infant sex were also analyzed in relation to cluster and tertile proportions. Maternal age was evaluated using beta regressions, consistent with the approach used for gestational age at delivery. Infant sex was evaluated using wilcoxon rank-sum tests from the “stat_compare_means” function from the “ggpubr” package (v0.6.0)^41^ in R.

## Results

### Image Segmentation Performance

Both image segmentation models achieved exemplary detection and segmentation performance at the patch and broader region of interest levels. At the patch level, the YOLOv8 and Mask R-CNN models achieved average precision scores of 0.801 and 0.768 (AP at an IoU threshold of 0.5) and 0.606 and 0.559 (AP at IoU thresholds from 0.5-0.95), respectively (**Table 2**). Furthermore, models employed within the inter-patch merging post-processing workflow demonstrated equally high-performing results, recovering original villi counts across regions spanning tens of thousands of pixels in either direction with an overall Pearson correlation of 0.98 and 0.94 across the test sets within the cross-validation folds, respectively (**Figure 1**). Based on these metrics, the YOLOv8 model was selected as the favored model. The model trained on the first fold was applied across all WSIs, resulting in 9,245,917 villi annotations.

**Figure 1:**
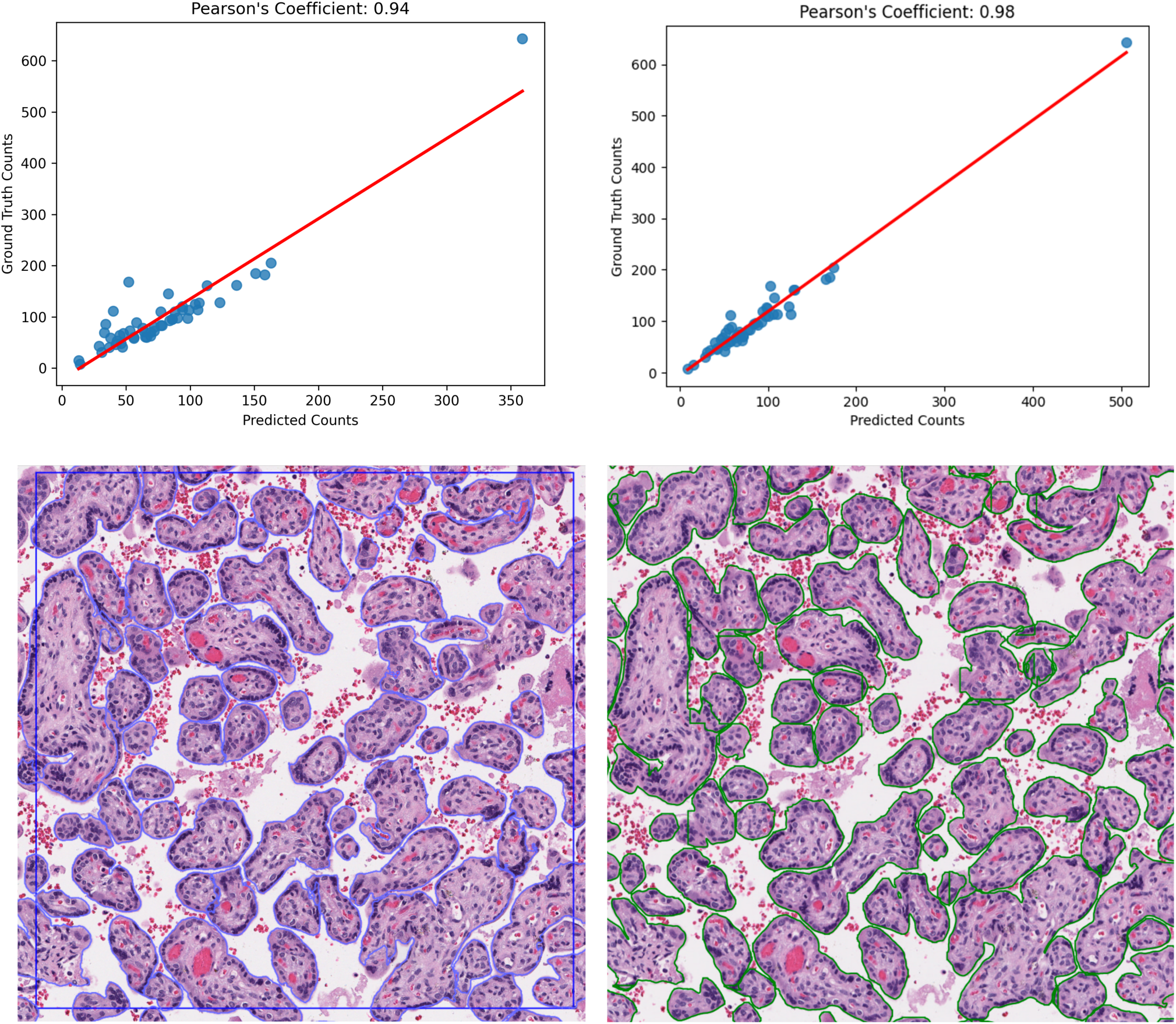
Correlation and Visualization of Predicted vs. Ground Truth Annotations **Panel A** displays scatterplots demonstrating the correlation between predicted counts and ground truth counts for the Mask-RCNN algorithm (left) and the YOLOv8 algorithm (right). The Pearson correlation coefficients are 0.94 and 0.98, respectively. **Panel B** is a sample from one ground-truth, hand-annotated WSI, and the YOLOv8 prediction of the same area. The left image displays hand annotations with additional support from SAM in blue outlines, and the right image shows predicted annotations from YOLO in green outlines, illustrating the differences and similarities between manual and automated annotation approaches.

**Table 2:** Model Performance for Mask-RCNN and YOLOv8.

|  | Mask R-CNN |  | YOLOv8 |  |
| --- | --- | --- | --- | --- |
|  | AP (IoU 0.5:0.95)* | AP (IoU 0.5) | AP (IoU 0.5:0.95) | AP (IoU 0.5) |
| Fold 1 | 0.608 | 0.801 | 0.654 | 0.820 |
| Fold 2 | 0.587 | 0.779 | 0.632 | 0.800 |
| Fold 3 | 0.575 | 0.783 | 0.627 | 0.817 |
| Fold 4 | 0.508 | 0.732 | 0.557 | 0.772 |
| Fold 5 | 0.519 | 0.745 | 0.562 | 0.795 |
| **Average** | **0.559** | **0.768** | **0.606** | **0.801** |

\*AP (IoU 0.5:0.95) is the average precision with different thresholds of Intersection over Union (IoU), ranging from 0.5 to 0.95, in increments of 0.05.
\*\*AP (IoU 0.5) is the average precision when evaluated at a threshold of 0.5.

### K-Means Cluster Proportions

K-means clustering was performed on a sample subset of n=150 annotated villi from each WSI (an aggregate of 222,319 villi) (**Figure 2)**. Upon visual inspection of the k=5 clusters, Clusters 2 and 3 were determined to be artifacts of the YOLOv8 segmentation model and not represent true villi. The k-means algorithm was then applied across all detected villi, then villi assigned to the artifact-labeled clusters (n=1,906,067) and any remaining villi with strong edges (n=338,224) were removed. 7,001,626 villi remained from clusters 1, 4, and 5. The primary difference between the remaining clusters was size, with Cluster 4 reflecting the smallest, most circular villi, Cluster 1 reflecting medium-sized villi, and Cluster 5 reflecting the largest, least circular villi (**Figure 2**).

**Figure 2:**
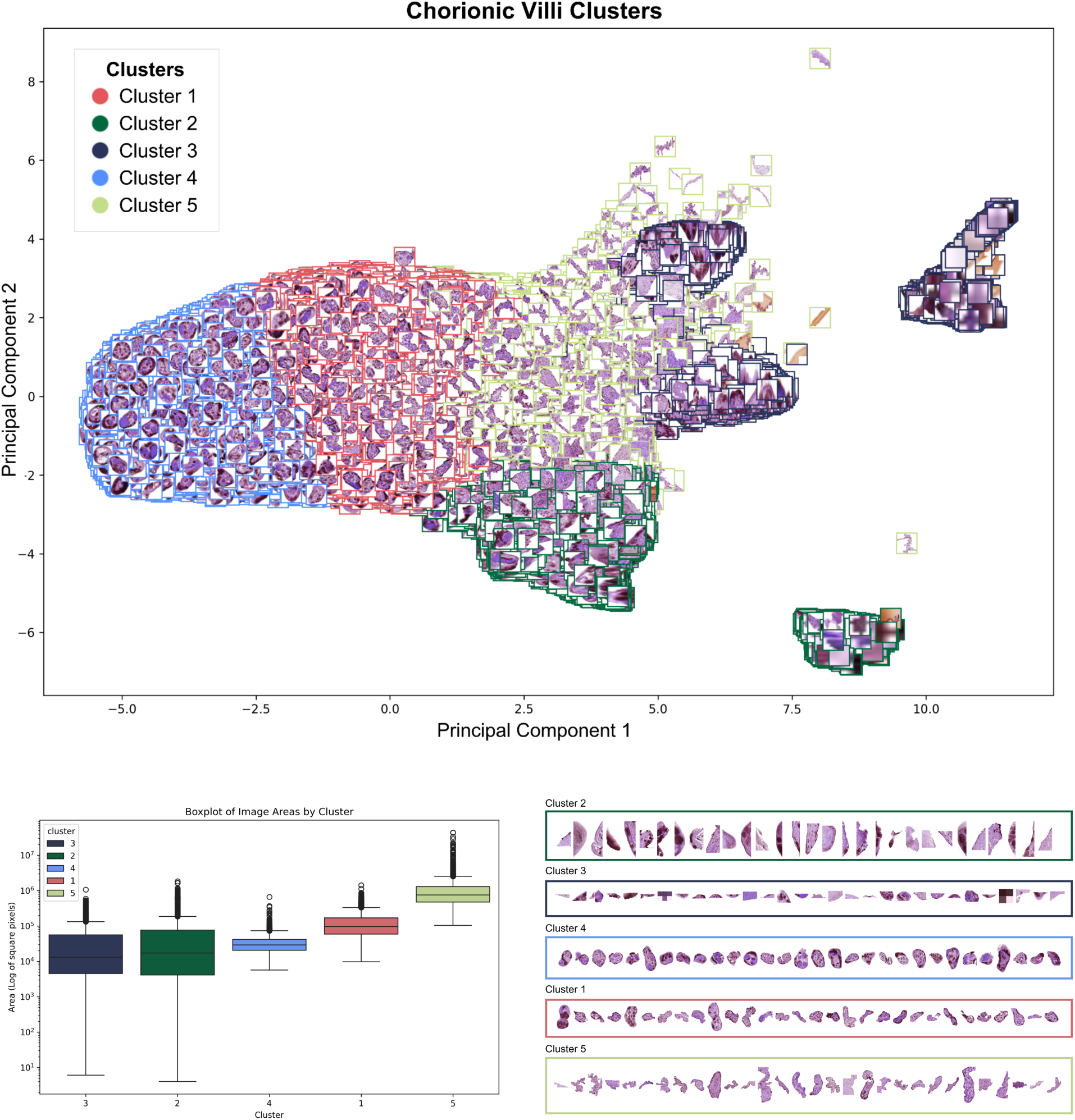
K-Means Clustering of Chorionic Villi This plot shows the results of the K-means clustering analysis. **Panel A** presents a scatterplot of the first two principal components (PC1 and PC2) with sampled images of villi colored by their respective clusters. There are five distinct clusters, 1-5. **Panel B** displays a boxplot of the log-transformed villi area (in square pixels) for each cluster. The median area and circularity for Cluster 4, 1 and 5, respectively are ∼3.5e4 pixels^2^, ∼0.77; ∼1.0e5 pixels^2^, ∼0.54; ∼5.10e5 pixels^2^, ∼0.28. **Panel C** provides a visual sample of 30 villi from each cluster and demonstrates the morphological variation within and between clusters.

**Figure S4** presents a comparison of the observed clusters and the reported range in size of villi subtypes from past studies. We found that the IQRs of villi size for Clusters 4, 1, and 5 align with expected ranges reported for terminal, mature intermediate, and immature intermediate villi, respectively.^7,10–12,21,42–46^ Further, **Figure S5** presents a bar chart that compares the proportion of all detected villi that were assigned to each specific cluster to the expected proportion of villi in each cluster based on prior reports.^6,10–14^ Compared to prior literature, this cohort shows fewer terminal villi (41.6% vs. ∼55.6%), and more mature (48.7% vs. ∼38.9%) and immature intermediate villi (9.7% vs. ∼5.6%).^6,10–14^ **Figure S6** presents the results of the K-Means Clustering algorithm for k=4 and k=6, the findings of which were compared with k=5 and the k=5 cluster model demonstrated the highest alignment with these established classifications of villi subtypes. Given the similarities by size and prevalence, Clusters 4, 1, and 5 will be referred to by these subtype classifications (i.e., terminal, mature, and immature) hereafter.

### Villi Area and Circularity Tertile Proportions

Tertiles of villi area and circularity were derived from global values of villi geometry across all WSIs to establish a simple biomarker of placenta growth. A general trend was observed with larger villi typically exhibiting more irregular shapes (i.e., lower circularity ratios) and smaller villi exhibiting more circularity (i.e., higher circularity ratios) (**Figure S7**). Nine tertile combinations were identified, with the highest relative proportions observed for S3:C1 (large, irregular villi) at 24% and S1:C3 (small, circular villi) at 22% (**Figure S8**).

### Placental Histopathologic Associations with Maternal and Infant Characteristics

Gestational age at delivery was statistically significantly associated with villi subtype proportions (p < 0.05) (**Figure 3**). Most notably, as gestational age increased, the proportion of mature intermediate villi increased (β=0.0093, 95% CI: [0.0032,0.015], p < 0.001). Proportions of terminal villi decreased modestly through gestation (β=-0.0042, 95% CI: [-0.015, 0.0063], p =0.059).

**Figure 3:**
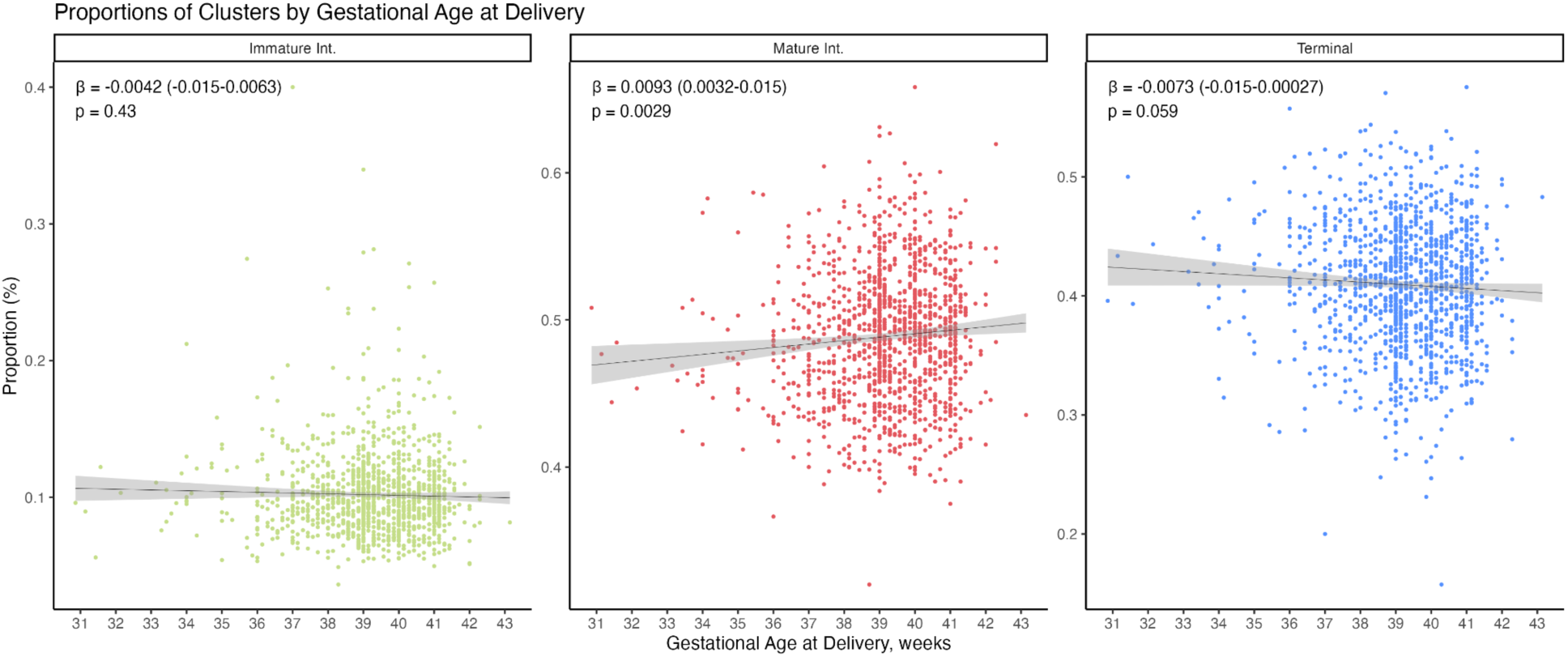
Gestational Age at Delivery and Cluster Proportion Line plots showing the relationship between gestational age (weeks) on the x-axis and the proportion of each cluster on the y-axis. Facets are grouped by cluster type (immature, mature, and terminal). The sum of the proportions across the three clusters for each WSI equals 1. P-values, beta estimates, and 95% confidence intervals are derived from a beta regression, where the proportion is the outcome and gestational age is the continuous predictor.

Gestational age at delivery was also assessed in relation to tertile proportions of villi size and circularity (**Figure 4**). Briefly, increasing gestational age at delivery was associated with a decrease in the proportion of the smallest, most circular villi (S1:C3) (β= -0.021, 95% CI: [-0.03, 0.011], p<0.001). Increasing gestational age at delivery was associated with a modest, but not statistically significant increase in the largest, most irregularly shaped villi (S3:C1) (β=0.0066, 95% CI: [-0.0014, 0.015], p = 0.11). In addition to these results, the change in proportion of S3:C1 (larger, irregular villi) was directly compared to S1:C3 (smaller, rounder villi) with a binomial logistic regression. We found that the probability of S1:C3 villi decreases compared to S3: C1 villi (β=-0.028, 95% CI: [-0.0295,-0.0265], p < 0.001).

**Figure 4:**
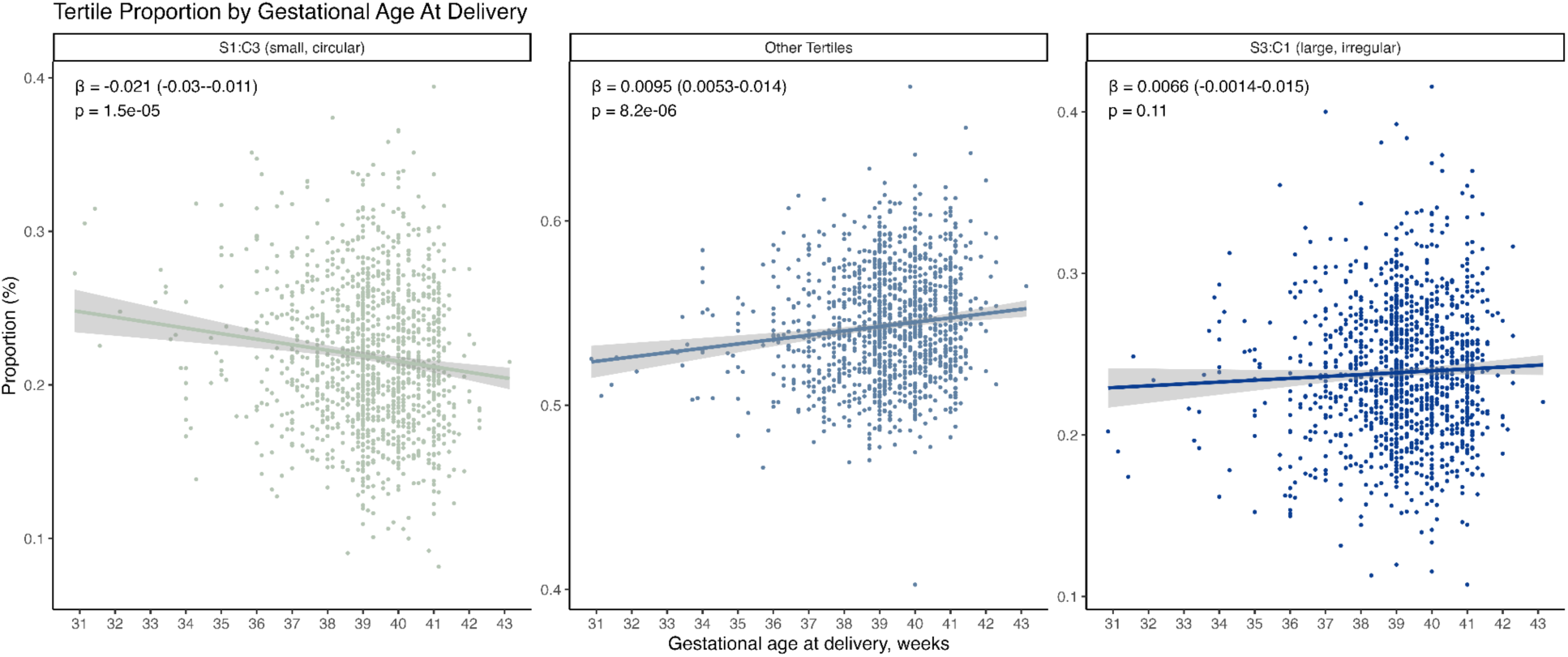
Gestational Age at Delivery and Area/Circularity Tertile Proportion Each villi was categorized into a size quartile representing the area (“S”) and a circularity ratio tertile (“C”) based on global total size and circularity ratio values. Higher “S” tertiles are larger villi (e.g. S3) and higher C tertiles are the roundest villi (e.g. C3). Thus, villi in the S1:C3 classification are the smallest and most round, and villi in the S3:C1 classification are the largest and most irregularly shaped. The proportion of villi in each WSI that belonged to each quartile and circularity tertile combination were calculated. Line plots showing the relationship between gestational age (weeks) on the x-axis and the proportion of each tertile on the y-axis. Facets are grouped by tertile combinations (S1:C3, S3:C1, and Other). The sum of the proportions across the three facets for each WSI equals 1. P-values, beta estimates, and 95% confidence intervals are derived from a beta regression, where the proportion is the outcome and gestational age is the continuous predictor.

A secondary analysis to examine temporal trends in villi classification, stratified by preterm and at term deliveries, revealed a significant increase in the log-odds ratio of a villus being classified as mature intermediate with each advancing week. This trend was observed in both preterm (β = 0.012; 95% CI: [0.008, 0.016], p < 0.001) and term deliveries (β = 0.005; 95% CI: [0.004, 0.007], p < 0.001) (**Figure S9, Table S3**). The interaction term (β=0.007; 95% CI: [0.002, 0.011], p = 0.003) suggested that the positive association between proportion of mature villi and gestational age was more pronounced in preterm deliveries. For immature intermediate villi compared to terminal villi, the log-odds ratio of a villi being classified as immature was not statistically significantly related to gestational age continuously for term deliveries, but it was for preterm deliveries (β=-0.010; 95% CI: [0.004, 0.017], p = 0.002) and increases in immature villi by gestational age was more pronounced in preterm deliveries (β=0.010; 95% CI: [0.003, 0.017]).

Maternal age and infant sex were not statistically significantly associated with any evaluated metric of villi quantification (**Figures S9-10**).

## Discussion

Objective measures of abnormal placental maturation may offer valuable insights into maternal and child health but remain limited by the accessibility and precision of histopathological examination. This study aimed to address these limitations through (1) development of an automated deep learning segmentation approach that is capable of categorizing placental chorionic villi, (2) quantification of the distribution of relevant villi subtypes and geometry, and (3) examination of these in the context of maternal and infant characteristics, including gestational age at delivery, maternal age, and infant sex. The successful application of an AI model on this large set of WSIs offers a promising approach to overcoming challenges in placental pathology, including time-consuming manual annotation and qualitative variability.

Our deep learning model achieved impressive accuracy, likely due to several key factors. Firstly, the training dataset was substantial in size (50 unique WSIs, ∼5,000 annotated villi,) and each villi was reviewed at least twice to improve data quality. Anecdotally, we found that the SAM plugin helped facilitate rapid curation of villi annotations where manual annotation proved cumbersome. Secondly, the integration of advanced deep learning architectures further enhanced the speed and accuracy of villi detection, as compared to manual annotation and previous models.^28,29^ Finally, the combination of a quality training dataset and state-of-the-art segmentation algorithms also ensured that the results were not subject to substantial error from additional post-processing, which is often needed when using models that rely strictly on boundary detection, color thresholding, and other image heuristics.

Downstream clustering effectively categorized chorionic villi into biologically relevant groups, including terminal, mature intermediate, and immature intermediate villi, consistent with prior literature that delineates these subtypes based on size and prevalence. This work reinforces the established understanding of villous structure while serving to validate the significance of our approach in enhancing the precision of villi characterization.

Based on the prior literature, we expected significant associations between villous surface area and gestational age. Many studies report increasing gestation to be associated with an increase in the proportion of terminal villi and a general decrease in size to facilitate better nutrient exchange.^21,26,31,47–49^ In contrast, we found that increasing gestational age is associated with an increase in mature intermediate and immature intermediate subtype proportions, relative to terminal villi. Similarly, when examining tertiles of area and circularity, we saw an increase in larger, more irregular villi (S3:C1) and a decrease in the overall proportion of small, circular villi (S1:C3) through gestation. Interestingly, some prior literature supports this finding and notes a general increase in villous surface area with increasing gestational age. For instance, Boyd et al.^20^ reported villous surface area increasing from 0.8 to 11.8 m² through gestation, while Jackson et al. (1992)^21^ reported an increase from 1.3 to 10.0 m². Jauniaux et al. reported that between 6-15 weeks of gestation, the volume fraction of villi increased from 2.67 to 4.11%.^31^ Within this context, several explanations for our finding are possible. First, our study included intermediate villi, whereas prior studies often focused on terminal villi alone. By incorporating a broader spectrum of villi, we captured growth patterns that may not have been emphasized previously. Second, our use of digital segmentation provides different measures from manual or semi-quantitative methods used historically, and are more comprehensive. Third, this dataset includes only placentas collected at delivery and doesn’t examine placentas from early and mid-gestation, highlighting the need for future studies that apply these methods to a broader gestational age sampling. Lastly, it’s biologically plausible that as gestation advances, individual villi have more time to grow and enlarge, contributing to the increased proportion of larger villi we observed.

While increasing gestational age is mostly associated with a decrease in villi size, categorical preterm birth has previously been associated with both accelerated and delayed villous maturation.^50,51^ Using statistical models that evaluated the interaction of preterm birth and gestational age, we noted that changes in the proportion of mature intermediate villi, relative to terminal villi, were more pronounced for preterm deliveries than for term deliveries, implying a slowdown in placental maturation after 37 weeks of gestation and suggesting that placental maturation is disrupted in preterm deliveries.

Finally, we were surprised to find that maternal age did not exhibit statistically significant associations with any measures of villi subtype or geometry in this study. In a study conducted by Žigić et al., authors found the total volume of terminal villi were significantly lower in an older pregnancy group compared to a younger group, which was not replicated in our study.^18^ However, we did not expect to find associations by infant sex; Jackson et al. reported no significant effects of infant sex on villous growth, which aligns with our results.^21^

Despite its strengths, there are various limitations. First, while YOLOv8 is fast and accurate, its computational demands may limit clinical use in locations with limited resources. Additionally, given the large number of available models (e.g., UNI, RT-DETR, and U-Net^52^), we focused on a few promising models since a comprehensive comparison was beyond the scope of our study and our focus was on the correlation of histopathologic features with maternal and infant characteristics using methods that were accessible and familiar to readers and easy to interpret for clarity and accessibility. In addition to examining other models, future work could examine other approaches and techniques such as model quantization, kernel fusion, compilation to lower-level languages, using foundation models specifically designed for pathology, considering non-Euclidean villous geometry, and knowledge distillation to smaller models. Finally, the algorithm might not be sensitive to unique developmental histopathological variations such as villous agglomeration or fibrin accumulation in the intervillous space, both of which are associated with villous maturation.^53^ These features could mistakenly be included in villi annotation contours, potentially inflating villi size in late gestation, therefore future research should better account for these maturational features.

This work has several strengths. First, the large sample size (1,510 WSIs and >9 million annotated villi) enables comprehensive characterization of villi across diverse morphologies. Second, the use of automated villi detection and segmentation algorithms reduces reliance on manual annotation, minimizes variability, and provides a standardized, objective quantification of chorionic villi in placentas collected at delivery. Developing a user-friendly villi quantification tool for pathologists could facilitate the integration of advanced imaging analysis into clinical practice, improving placental health assessments in diverse settings.

In conclusion, this study demonstrates the potential of automated deep learning algorithms to enhance placental chorionic villi characterization, offering insights into placental development and function. The precise quantification of villi subtypes and geometry reveals significant associations with gestational age at delivery. While this objective approach to villous assessment is promising, further studies across a broader range of gestational ages and villous subtypes are needed. The advancement and integration of automated tools into routine placental pathology could improve clinical practices, accelerate placental evaluations, enhance the diagnosis of abnormalities, and ultimately contribute to better maternal and fetal health outcomes.

## Supporting information

Graphical Abstract

Highlights

## Data Availability

All data produced in the present study are available upon reasonable request to the authors

## Acknowledgements

MK is the Principal Investigator of the New Hampshire Birth Cohort Study and designed and oversaw the original cohort data and sample collection, including placental biopsies. TP facilitated specimen transport and coordination between the NHBCS and Pathology Shared Resource. HW performed research scanning. JB performed histopathologic assessment and supervised JC, AS, NR, CJ, ECA, SP, ZCB, AC, OO, AK, and LH for annotation tasks. CH contributed to conceptualization of the project with ECA, JJL, and JLB, provided feedback on the analysis plan, helped supervised statistical analyses conducted by ECA with JJL, and reviewed and edited the manuscript. GS, ECA and CJ implemented segmentation approaches. EZ and LZ implemented clustering approaches. EZ and CJ implemented post-processing techniques, refined by ECA. ECA performed statistical analyses leveraging the NHBCS metadata and villi quantifications and drafted the initial manuscript.

## Competing Interests

None to declare.

## Ethics Statement and Patient Consent

The study was approved by the Committee for the Protection of Human Subjects at Dartmouth College (CPHS# STUDY00020844). Written informed consent was obtained from all subjects involved in the study prior to engagement in any study activities.

## Funding

This study was funded by the National Institutes of Health [P20GM104416, P20GM130454, P01ES022832, UG3OD023275, UH3OD02327] and the Burroughs-Wellcome Fund Big Data in the Life Sciences training grant at Dartmouth.

## Abbreviations

NHBCS: The New Hampshire Birth Cohort Study
SAM: Segment Anything Model
WSI: Whole Slide Image

## Notes

### Competing Interest Statement

The authors have declared no competing interest.

### Funding Statement

This study was funded by the National Institutes of Health [P20GM104416, P20GM130454, P01ES022832, UG3OD023275, UH3OD023275] and the Burroughs-Wellcome Fund Big Data in the Life Sciences training grant at Dartmouth.

