## Supplementary Materials for "Association of Deep Learning-Derived Histologic Features of Placental Chorionic Villi with Maternal and Infant Characteristics in the New Hampshire Birth Cohort Study"

### **Supplemental Figures and Tables**

##### Table S1: Previously reported chorionic villi metrics

|  | **Stem** | **Immature Intermediate** | **Mature Intermediate** | **Terminal** |
| --- | --- | --- | --- | --- |
| **Diameter** | 80-3,000 µm^1,2^ | 40-400 µm^1,3^ | 50-150 µm^1,3–5^ | 30-100 µm^3,5–10^ |
| **Proportion in term**^1,5,7,11–13^***** | 20-25% | 3-5% | 20-35% | 40-50% |

*References for supplemental tables and figures are included at the end of the supplemental materials

S2: Hand Annotated Sample Size vs. Full Cohort

| **Characteristic** | | **Not Annotated (n=1,482)** | **Annotated (n=49)*** | **P-value**** |
| --- | --- | --- | --- | --- |
| Gestational age, weeks | | 39.29 (1.86) | 39.86 (1.93) | 0.14 |
| % Preterm (< 37 weeks gestation at delivery) | | 1.00 (2%) | 98 (6.60%) | 0.26 |
| Maternal BMI, kg/m^2^ | | 24.89 (7.10) | 23.90 (5.77) | 0.31 |
| Placenta Weight, grams | | 580.00 (170.00) | 545.00 (125.00) | 0.43 |
| Birth weight, grams | | 3420.00 (636.50) | 3430.00 (495.50) | 0.56 |
| Noted placenta abnormalities | | 77 (5.2%) | 5 (10.2%) | 0.23 |
| Infant sex, male | | 631 (42.6%) | 23 (46.9%) | 0.66 |
| Enrollment year | 2013-2015 | 372 (25.1%) | 1 (2.0%) | 7.02e-27 |
|  | 2016-2018 | 334 (22.5%) | 47 (95.9%) |  |
|  | 2019-2022 | 650 (43.9%) | 0 (0.0%) |  |
|  | NA | 126 (8.5%) | 1 (2.0%) |  |
| Smoking | Smoked while pregnant | 4 (8.2%) | 63 (4.3%) | 4.39e-3 |
|  | Ever Smoked (but not during pregnancy) | 0 (0.0%) | 48 (3.2%) |  |
|  | Second-hand smoking exposure during pregnancy | 5 (10.2%) | 40 (2.7%) |  |
|  | No reported smoking or second-hand smoking exposure | 39 (79.6%) | 1090 (73.5%) |  |
|  | Missing | 0 (0.0%) | 115 (7.8%) |  |
| Delivery Type | Vaginal Delivery | 886 (59.8%) | 36 (73.5%) | 0.63 |
|  | C-section | 369 (24.9%) | 12 (24.5%) |  |
|  | NA | 0 (0.0%) | 0 (0.0%) |  |
| Parity | 0 | 577 (38.9%) | 20 (40.8%) | 0.68 |
|  | 1 | 490 (33.1%) | 17 (34.7%) |  |
|  | 2 | 167 (11.3%) | 8 (16.3%) |  |
|  | ≥3 | 56 (3.8%) | 3 (6.1%) |  |

Values are in the format of median (IQR) or N (%)

*Although 50 WSIs were hand-annotated and used for training, one of these WSIs was later identified as not belonging to an NHBCS participant

**Statistical comparisons for continuous variables used the Wilcoxon Rank Sum test, and Fisher’s Exact Test was used for categorical variables

##### Figure S1: Conceptual Diagram of Placental Villi Maturation

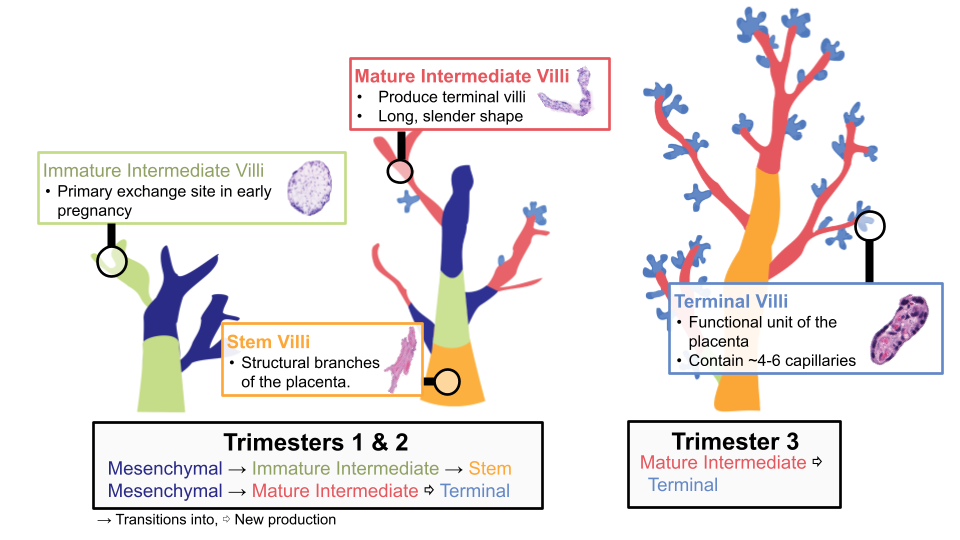

Normal placental maturation begins with the growth of mesenchymal villi, which differentiate into either immature intermediate or mature intermediate villi, representing key developmental trajectories. During the second trimester, immature intermediate villi develop into stem villi, which are the largest villi that connect the placental vasculature to the umbilical cord. At the end of the second trimester and through the third trimester, mature intermediate villi produce terminal villi through non-branching angiogenesis, which are the smallest villi and generally round or oval with 4-6 capillaries in each villi.

##### Figure S2: Whole Slide Image (WSI) Sample Selection

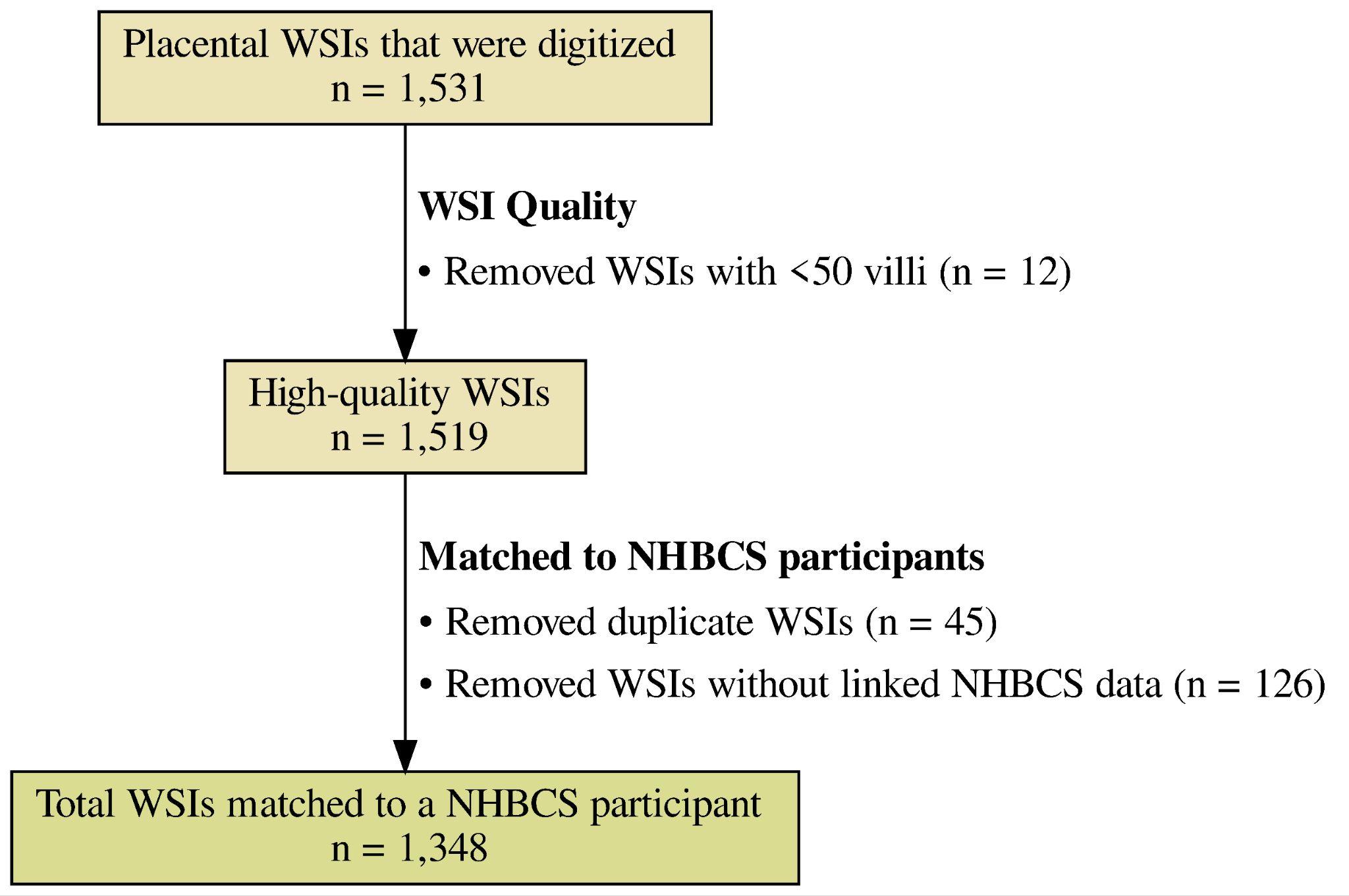

This flowchart illustrates the process of selecting which placental WSIs to use for the primary analysis. This took part in two stages: first, ensuring high-quality WSIs that had a sufficient number of villi to examine, and second, matching scanned WSIs to NHBCS participants.

N=12 WSIs were removed because they had less than 50 villi, n=45 were removed because they matched a duplicated NHBCS participant, and n=126 were removed because they could not be linked to an NHBCS participant

##### Figure S3: Missingness overlap for gestational age at delivery, maternal age, and infant sex

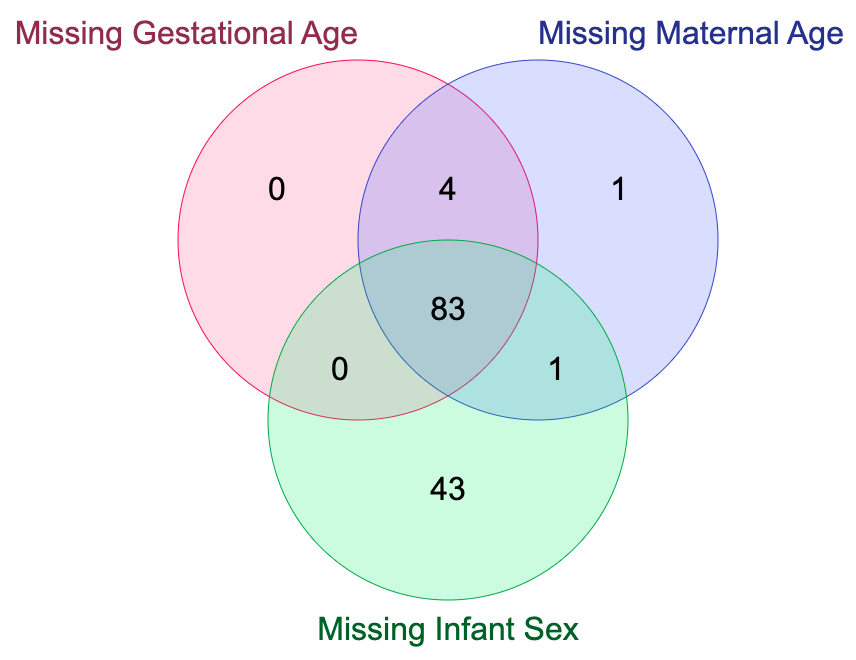

In each analysis, some participants were missing maternal or infant characteristic data. This venn diagram illustrates the overlap of participants who were missing information. Of the total sample size, 89 participants had missing gestational age at delivery data, 87 had missing maternal age data, and 127 had missing infant sex data. Of the 132 participants missing any information, 83 (62.9%) were missing all three maternal and infant characteristics.

##### Figure S4: Boxplot of cluster size and reported ranges of size for villi types

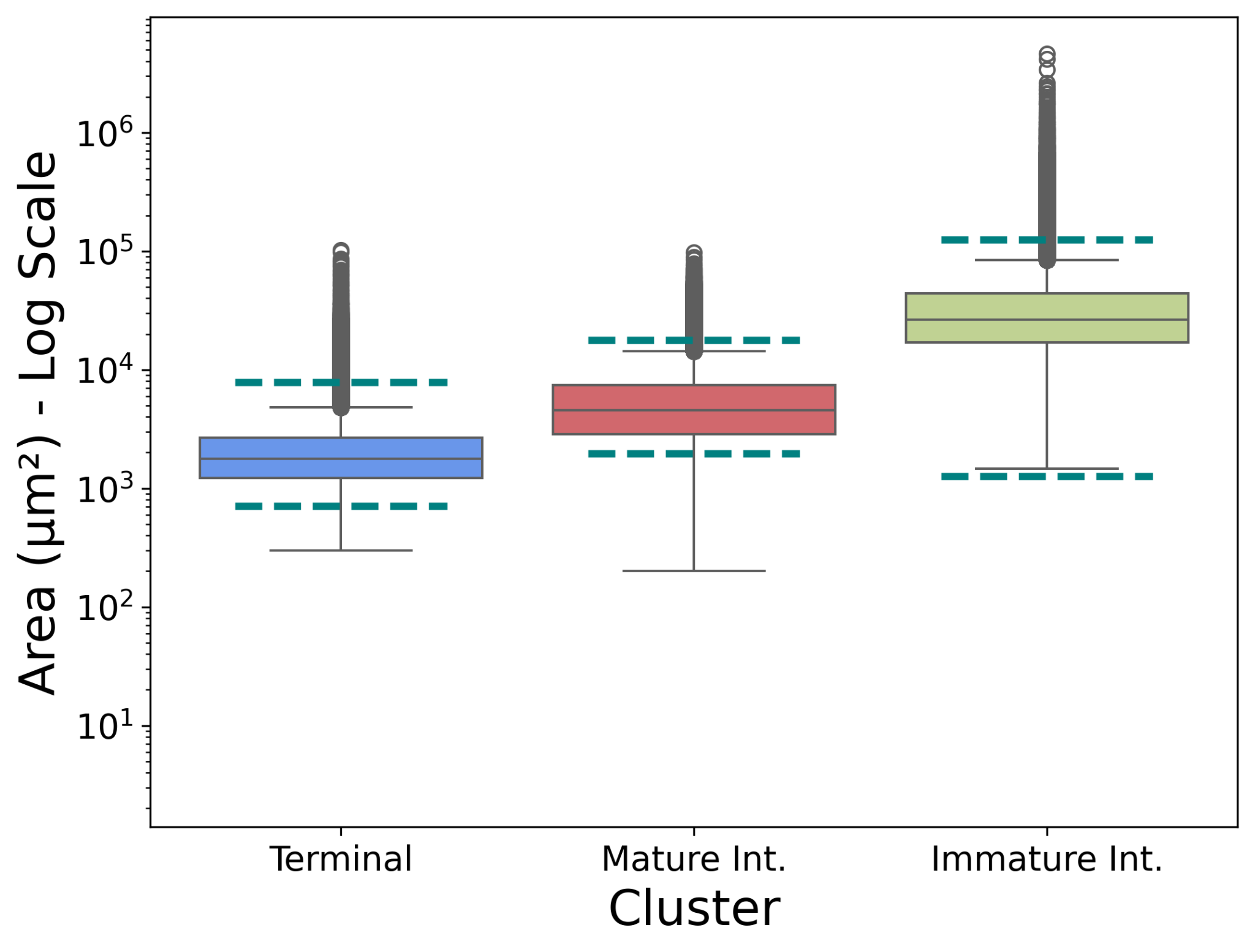

This boxplot illustrates the areas of all villi, grouped by cluster. Different types of villi in term placentas—Terminal, Mature Intermediate, and Immature Intermediate—are known to vary in size. Reported diameters for these villi types range from 40–400 µm for Immature Intermediate, 50–150 µm for Mature Intermediate, and 30–100 µm for Terminal villi. These diameters (µm) were converted into areas (µm²) for comparison with the clustered villi data. In our dataset, Terminal villi (Cluster 4) have an IQR of 1,227 – 2670 µm²; compared to the broader reported range of 707 – 7,854 µm². Mature Intermediate villi (Cluster 1) have an IQR of 2,861 – 7,425 µm²; with the broader reported range from 1,964 – 17,672 µm². Immature intermediate villi (Cluster 5) have an IQR of 16,994 – 43,955 µm²; with a broader reported range from 1,257 – 125,664 µm².

##### Figure S5: Stacked bar chart comparing villi type proportions

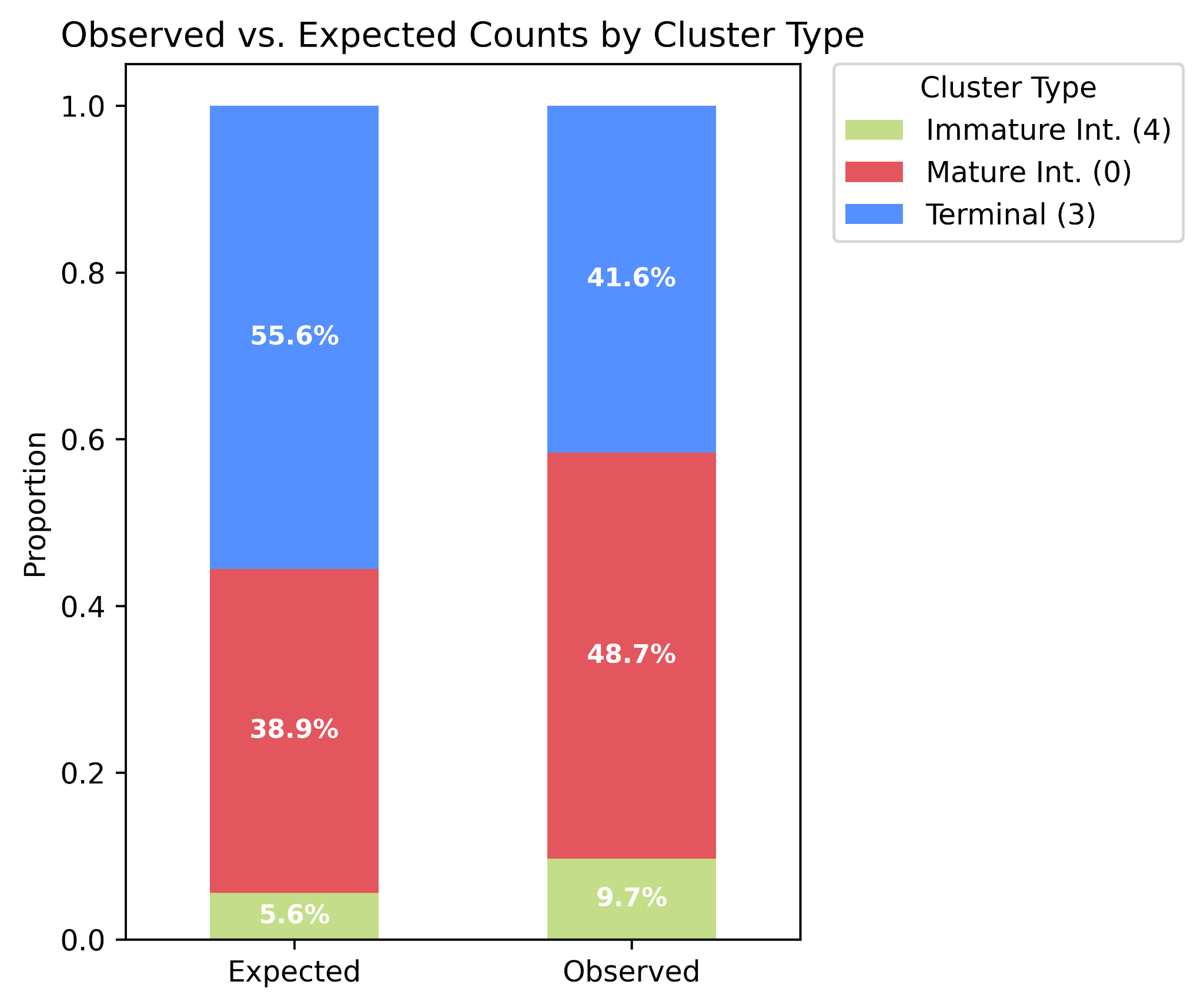

This stacked bar chart presents a comparison of the expected villi distribution, as reported in the existing literature, with the observed villi proportions derived from the K-means clustering algorithm.

##### Figure S6: Clustering algorithm for K=4 and K=6

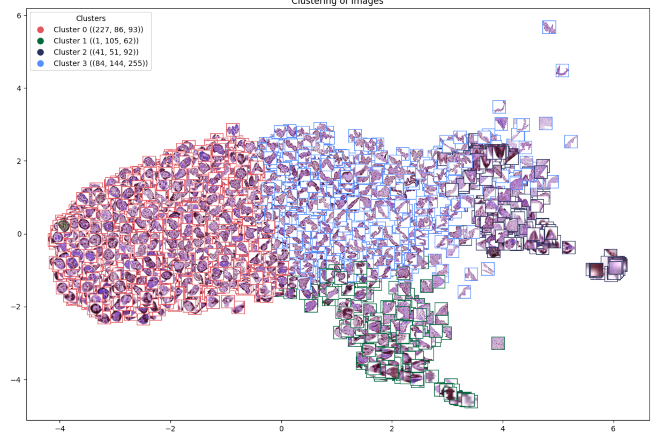

This plot shows the results for the k=4 (Panel A) and k=6 (Panel B) K-Means clustering algorithm. The two clusters that represent annotation artifacts (Cluster 2 and 3 in the k=5 clustering here) are consistent in these two plots. The k=4 model divides the same villi from the k=5 model Clusters 1, 4, and 5 into two groups instead of three. The k=6 model splits Cluster 1 from the k=5 model into two separate groups. Morphological and size evaluation of these two groups did not confer confidence in a unique and biologically relevant villi subtype.
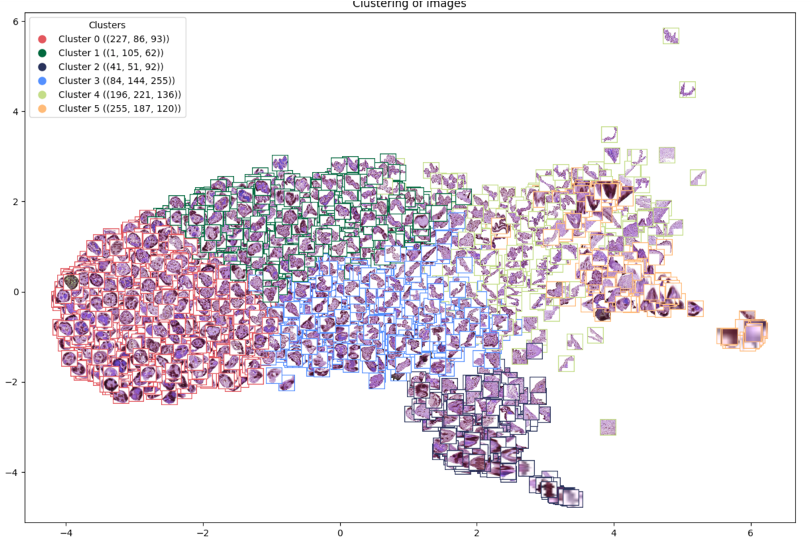

##### Figure S7: Area and Circularity of Villi

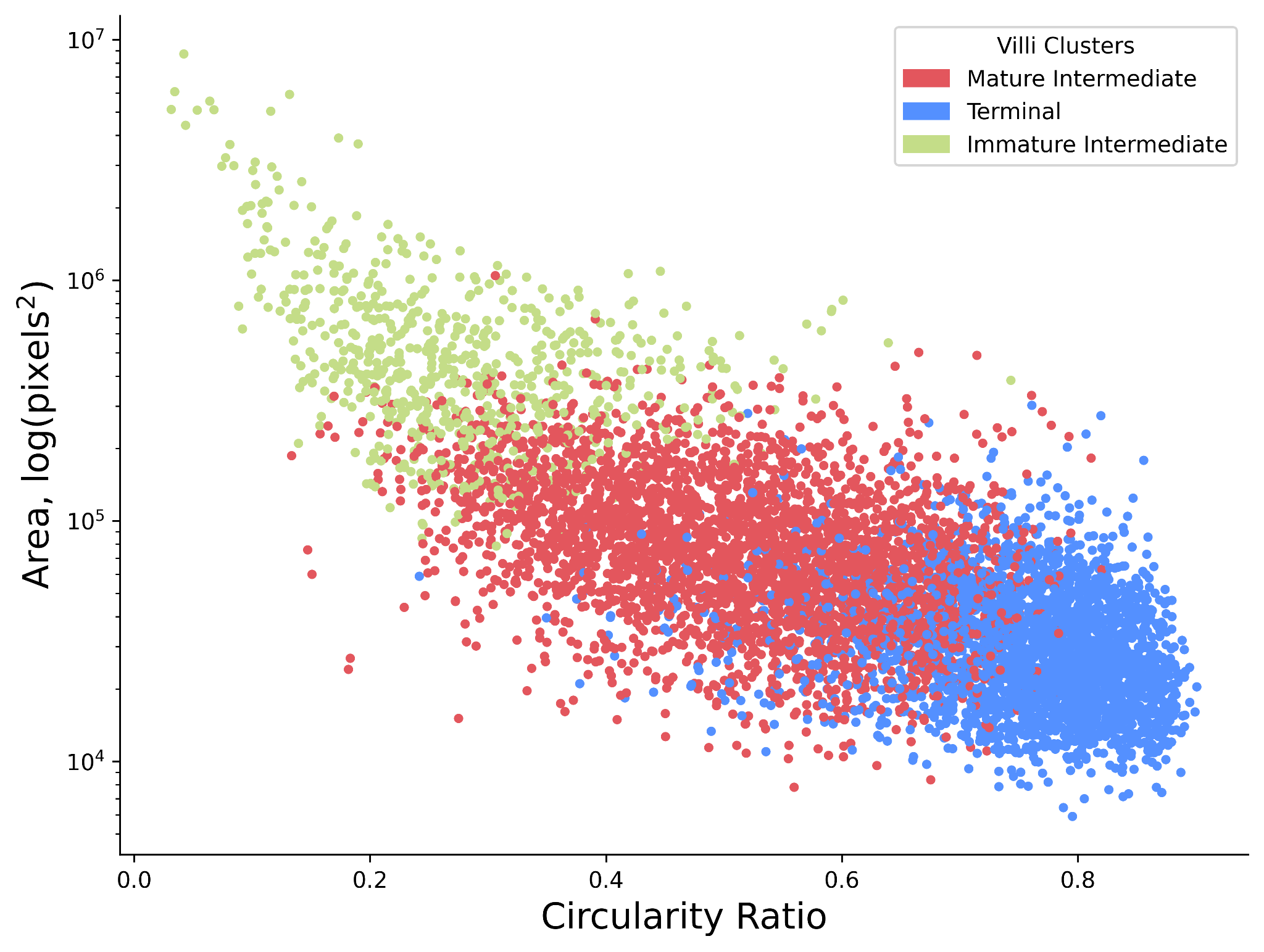

This plot represents a randomly selected sample of 7,002 villi, plotted by their area (y-axis) and circularity ratio (x-axis), with colors indicating their respective clusters. A general downward trend is observed, suggesting that as villi decrease in size, they tend to become more circular.

##### Figure S8: Heatmap of Area and Circularity Tertiles

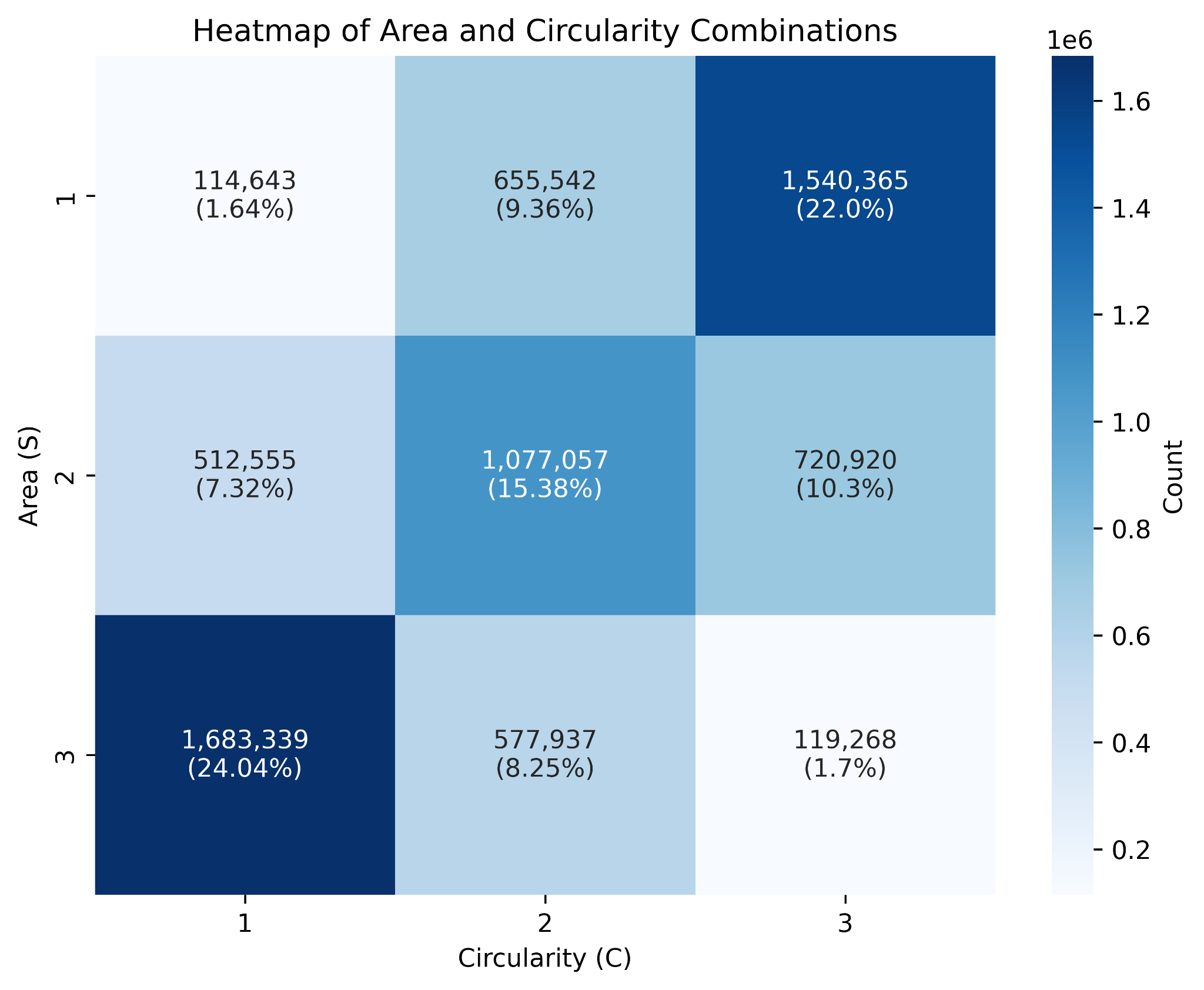

This heatmap shows the classification of all villi into tertile combinations of villi area (in pixels^2^) (“S”) and Circularity Ratio (“C”). The combinations with the highest proportion were S3:C1 and S1:C3 which represent that largest, most irregular villi and the smallest, most circular villi, respectively.

##### Figure S9: Gestational Age at Delivery, by Preterm Status

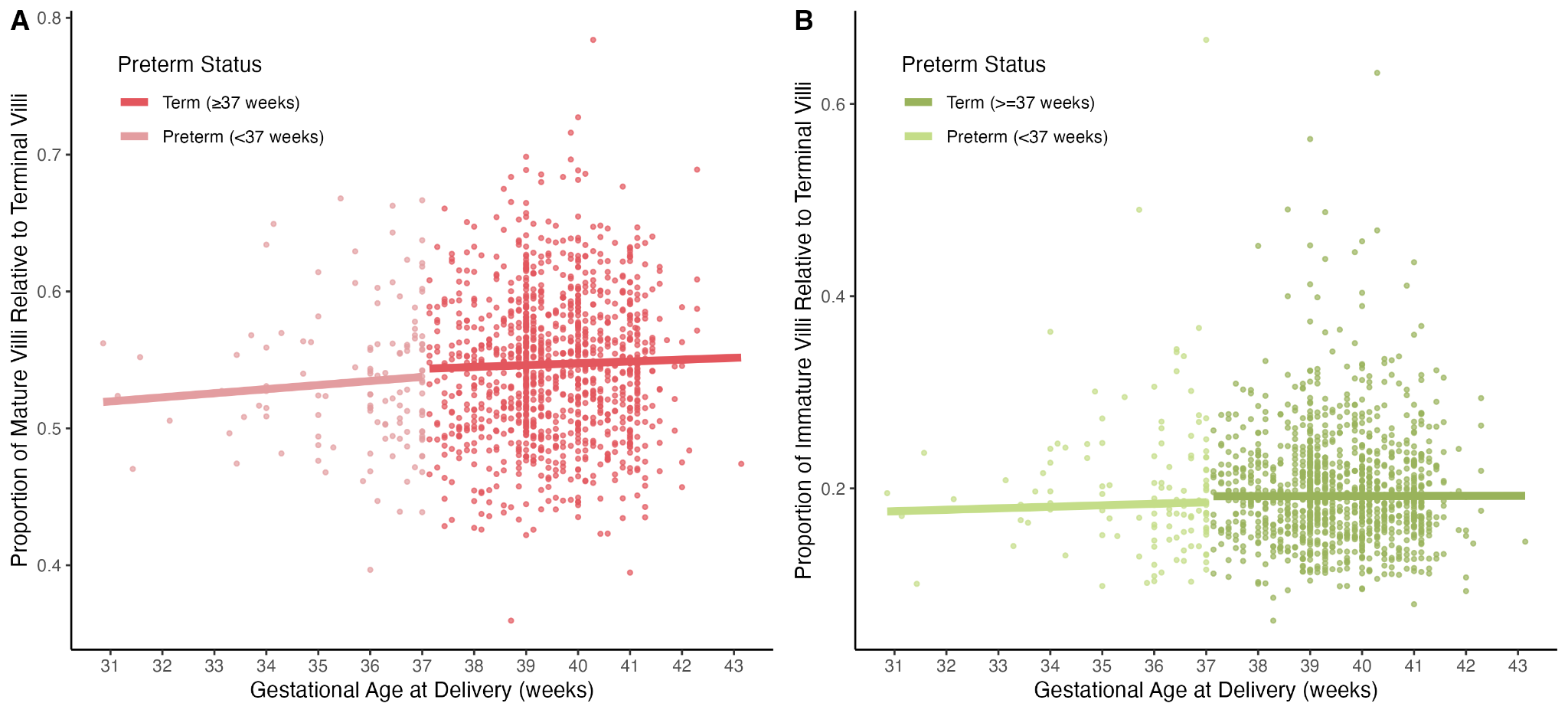

**Panel A** presents the proportion of mature intermediate villi relative to terminal villi, by gestational age and preterm status. **Panel B** presents the same comparison, but for immature intermediate villi relative to terminal villi.

Table S3: Logistic regression results for mature intermediate and immature intermediate proportions variations by gestational age at delivery and preterm status compared to the proportion of terminal proportions

|  | **Coefficient** | **β** | **95% Confidence Interval** | **P-Value** |
| --- | --- | --- | --- | --- |
| Mature Intermediate | Gestational age at delivery, weeks \| Not preterm (>= 37 weeks gestation) | 0.005 | (0.004, 0.007) | 0.000 |
|  | Gestational age at delivery, weeks given preterm (< 37 weeks gestation) | 0.012 | (0.008, 0.016) | 0.000 |
|  | Interaction between preterm and gestational age at delivery | 0.007 | (0.002, 0.011) | 0.003 |
| Immature Intermediate | Gestational age at delivery, weeks \| Not preterm (>= 37 weeks gestation) | 0.000 | (-0.002, 0.003) | 0.747 |
|  | Gestational age at delivery, weeks given preterm (< 37 weeks gestation) | 0.010 | (0.004, 0.017) | 0.002 |
|  | Interaction between preterm and gestational age at delivery | 0.010 | (0.003, 0.017) | 0.007 |

##### Figure S10: Maternal Age and Cluster Proportion/Tertile Proportion

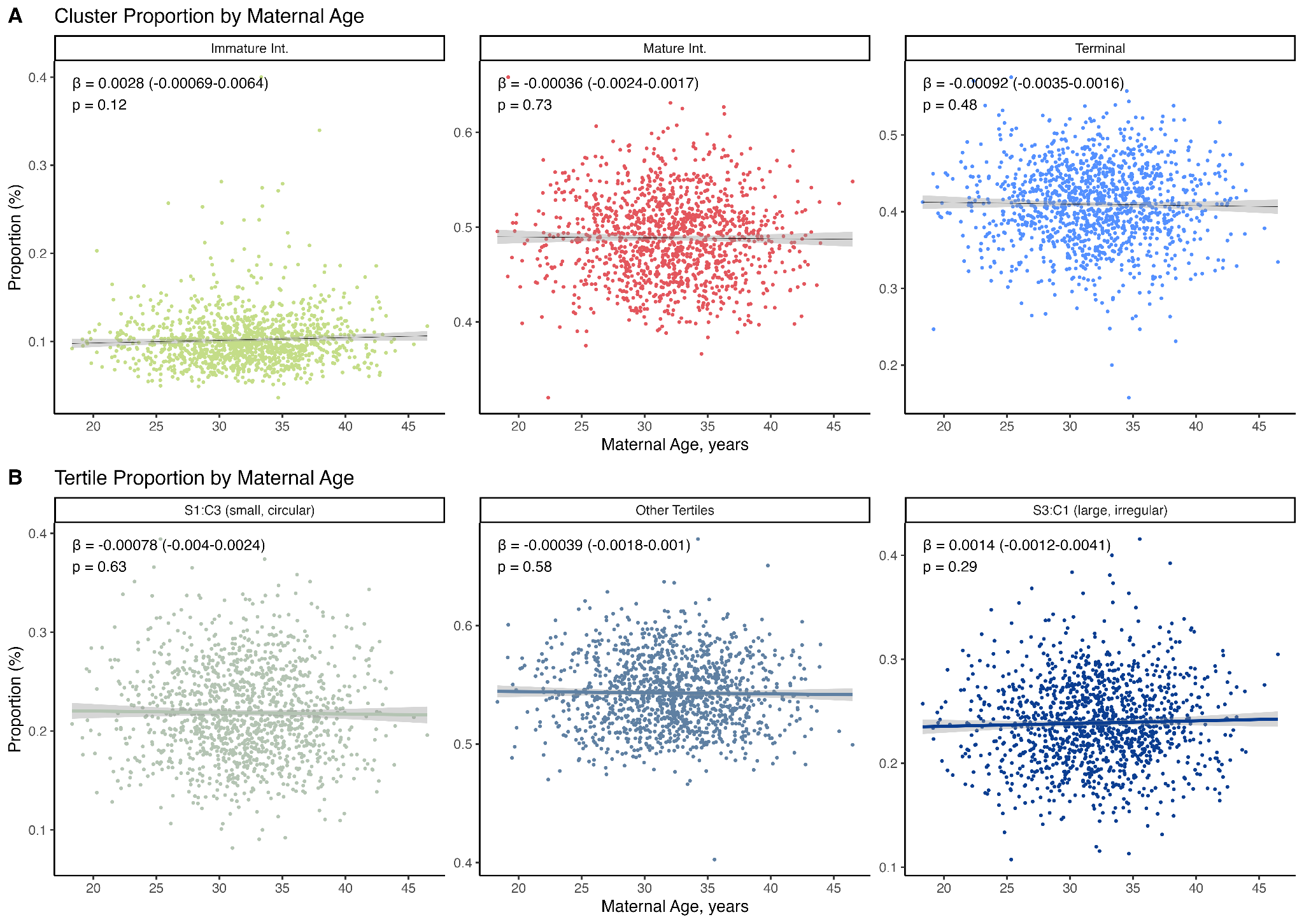

Line plots showing the association of maternal age with (1) Cluster proportion (**Panel A**) and (2) Tertile Proportion (**Panel B**). Effect estimates, 95% CIs, and p-values are provided given beta regressions. Maternal age is not statistically significantly associated with cluster or tertile proportion.

##### Figure S11: Infant Sex and Cluster Proportion/Tertile Proportion

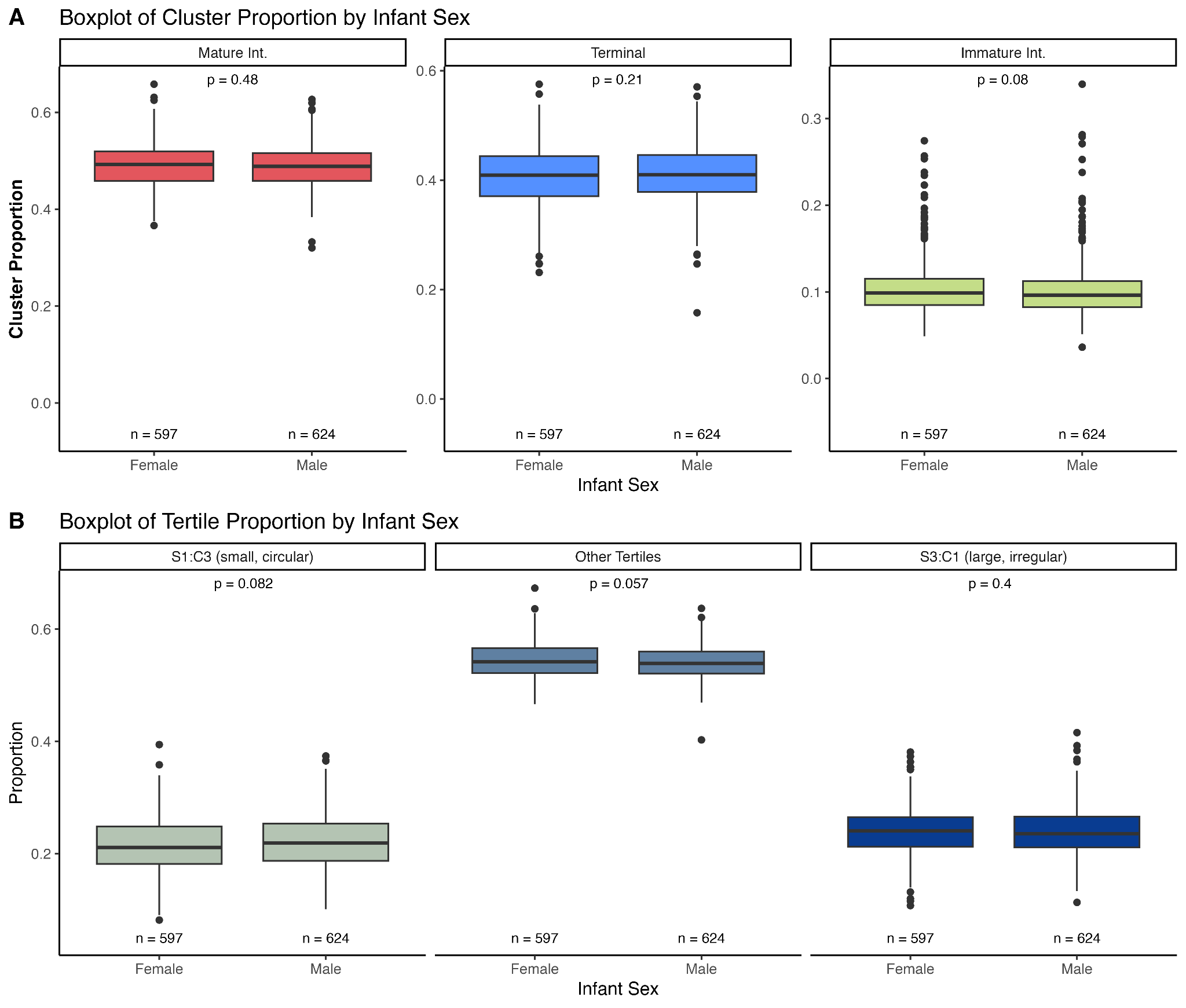

Boxplots showing the association of infant sex with (1) Cluster proportion (**Panel A**) and (2) Tertile Proportion (**Panel B**). P-values are provided from wilcoxon signed rank-sum tests (“wilcox.test” from R). Infant sex is not statistically significantly (p < 0.05) associated with cluster or tertile proportion.

##### **Supplemental Text**

##### WSI Annotation

WSIs from the digital scanning were annotated using QuPath, an open-source software for pathology image viewing.^14^ For each WSI, rectangular regions of interest (ROIs) were selected for contour outlining. Villi were initially annotated by a team of 10 individuals for further review and quality assessment by a board-certified gynecologic pathologist. Given the time and resource constraints of manual annotation of villous structures, most villi contours were made using the Segment Anything Model (SAM) plugin for QuPath, a semiautonomous annotation tool which accelerated the annotation process while maintaining precision.^15^ SAM is a prompt-guided deep learning-based segmentation tool– akin to “magic wand” tool commonly employed in image editing/annotation software– trained on over one billion segmentation masks across diverse objects and scenes, making it well suited to generate precise segmentation masks for new, complex structures such as villi based on user-defined input prompts (e.g., points, boxes, regions of interest). Using this plugin, annotators drew box prompts around candidate villi within the ROI, and the SAM QuPath plugin produced smooth annotations that tightly contoured each villous structure.

After the first round of annotations was completed, E.C.A. reviewed the annotations with J.B., a board-certified gynecologic pathologist. Given inherent individual variance in annotating villi, discrepancies in villi annotations were apparent between annotators. To address this, J.B. and E.C.A. established that a villi would be annotated if it met the following criteria: it was a region with at least one visible syncytiotrophoblast cell and one stromal cell or visible evidence of a capillary. Based on this standardization, E.C.A. reviewed all of the villi annotations to ensure they met the established criteria. Additionally, E.C.A. verified that the ROIs of each WSI were evenly divisible by 1024x1024 pixels in accordance with predefined image detection requirements and empirical performance analyses. In some cases, this required further villi annotation when ROI expansion was necessary to meet the pixel divisibility standard. J.B. then reviewed a sample of WSIs to confirm their validity and quality. In total, n=4,975 villi were annotated from 50 regions of interest that, in aggregate, covered 9.63 million pixels^2^. The participants characteristics from the 50 WSIs annotated were not statistically significantly different from the analytic subset (**Table S2**).

Training data for each model was produced using a non-overlapping sliding window approach with a square patch size of 1024x1024 pixels. The 50 annotated WSI ROIs were divided into five folds, with each fold containing 40 training WSI ROIs and 10 validation WSI ROIs. Patches from the same ROI were not split across training and testing folds. Across the five folds, the number of training patches were 732, 742, 746, 733, and 640, respectively, with corresponding testing patches of 166, 156, 152, 166, and 258.

##### Instance Segmentation Model Details

The YOLOv8 model was trained for 20 epochs (or until convergence) using the AdamW optimizer with a learning rate set to 2e-3 and a momentum of 0.9. A batch size of 8 was used. To diversify the training dataset, improve generalization, and reduce overfitting, image augmentations were used, including vertical flip (p = 0.5), horizontal flip (p = 0.5), rotation (90 degrees), translation (100 pixels in any direction), and random saturation (p = 0.5).

The Mask R-CNN model was trained for 10 epochs (or until convergence) using the SGD optimizer with a learning rate set to 1e-3 and a batch size of 2. The same image augmentations used on the training dataset for the YOLOv8 model were applied to this model.

To address truncation of villi at detection patch edges, we applied a sliding window approach with overlapping patches on held-out tissue slides, where each villi was often detected in multiple overlapping patches. Detections corresponding to the same villi were merged using a connected components graph, constructed based on whether the Intersection over Union (IoU) between adjacent instance masks exceeded 0.05—a threshold determined through sensitivity analysis which compared these thresholds using various villi and slide-level performance metrics.

All model performances were evaluated via five-fold cross-validation, and performance metrics were captured through segmentation mean average precision (mAP) at an intersection over union (IoU) threshold of 0.5 (mAP50), as well as the average mAP across varying IoU thresholds, running from 0.5 to 0.95 in increments of 0.05 (mAP50-95). The mAP at varying IoU thresholds ranges from 0 to 1, with values closer to 1 indicating better performance. We additionally validated the models by examining the correlation between the number of ground-truth annotated villi and the model’s predicted villi from the same ROIs. Concordance between the number of ground truth and predicted annotations was assessed using Pearson correlation.

The finetuned YOLOv8 model was applied to the rest of the WSIs. For each image, YOLOv8 detected and classified villi regions based on the learned features from the training model. The annotations for each WSI were saved in a geojson format. J.B. again reviewed a random sample of the annotated WSIs to confirm their validity and quality.

##### Villi Clustering

K-means clustering was performed on a random subset of n=150 annotated villi from each WSI (an aggregate of 222,319 villi) to separate the annotations into interpretable, biologically relevant groups. Two features were extracted for each villi: visual features from a pre-trained VGG16 convolutional neural network and the length and width of each villi. The VGG16 model from the Keras package (v3.6.0)^16^ was initialized with weights pre-trained on the ImageNet dataset, and the feature activations were taken from the penultimate fully connected layer ('fc1'), yielding a 4,096-dimensional feature vector for each villi. UMAP (v0.5.6)^17^ was applied for dimensionality reduction, using a minimum distance of 0.1, 15 neighbors, and 2 output components.

Clustering was performed using the k-means algorithm from the sklearn library (v1.3.2)^18^ in Python version 3.11.9.^19^ Cluster sizes, “k”, from 1-6 were evaluated and the final “k” was chosen based on visual inspection. The selected clustering model (including the UMAP projection) was saved and then applied to the rest of the villi across all the WSIs, yielding a cluster label for every villi. Upon visual inspection, some clusters were found to contain predominantly model artifacts, meaning villi annotations that were not true villi. These annotations lacked nuclei, capillary evidence, and/or synctiotrophoblastic cells, and often had straight perimeter lines instead of the normal villous contours. Any annotation within these groups were excluded from further analysis.

In addition to excluding villi that were clustered into artifact groups, a custom algorithm was developed to filter out any remaining annotation artifacts that were not true villi. Villi annotations were excluded if either (1) 40% or more of their total perimeter consisted of a straight line, or (2) the combined length of straight segments each exceeding 10% of the total perimeter accounted for more than 40% of the perimeter. The second criterion was primarily introduced to capture instances where villi annotations exhibited straight-edged corners that, individually, did not surpass the 40% threshold. Finally, after removal of all artifact villi, the proportion of villi in each cluster in each WSI was calculated by dividing the number of villi in each assigned cluster by the total number of villi in the WSI.

##### Tertile Classification Creation

The median size of villi in each cluster was found by taking the median area, in pixels^2^, of each villi polygon. Circularity ratios were calculated using custom code, which involved the following steps: finding the perimeter of the villi polygon, finding the area of a reference circle that has the same perimeter of the polygon, and then dividing the true area of the polygon by the reference circle. The range of circularity ratios was from 0 to 1, where ratios closer to 1 indicated a more circular polygon, while lower ratios indicated a more irregular shape
