## Supplementary material for "Association of Deep Learning-Derived Histologic Features of Placental Chorionic Villi with Maternal and Infant Characteristics in the New Hampshire Birth Cohort Study": Graphical Abstract

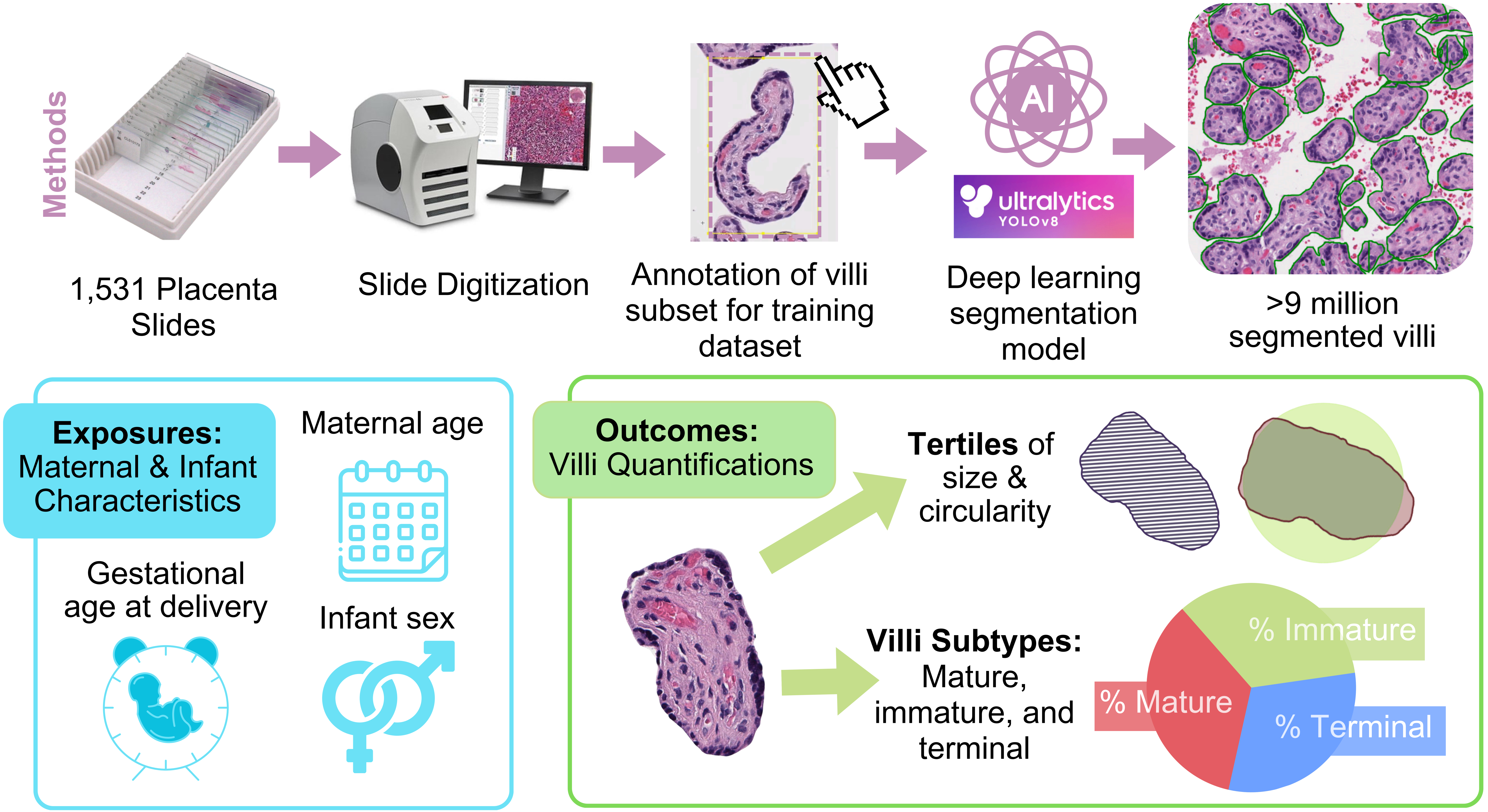


**Result**: Deep learning segmentation of placental villi revealed that increasing gestational age is associated with more mature intermediate villi and fewer small, circular villi
