## Supplementary material for "Association of Deep Learning-Derived Histologic Features of Placental Chorionic Villi with Maternal and Infant Characteristics in the New Hampshire Birth Cohort Study": Highlights

1. Objective measures of placental maturation are limited by subjective pathologic examination
2. Machine learning workflow detected 9M villi from 1,531 placental slide images
3. Unsupervised clustering identified distinct terminal and intermediate villi subtypes
4. Increasing gestational age is linked with higher proportion of intermediate villi
5. Automated placental analysis could improve assessment of maternal-fetal health outcomes
